# Multiplexed Assays of Variant Effect and Automated Patch-clamping Improve *KCNH2*-LQTS Variant Classification and Cardiac Event Risk Stratification

**DOI:** 10.1101/2024.02.01.24301443

**Authors:** Matthew J. O’Neill, Chai-Ann Ng, Takanori Aizawa, Luca Sala, Sahej Bains, Annika Winbo, Rizwan Ullah, Qianyi Shen, Chek-Ying Tan, Krystian Kozek, Loren R. Vanags, Devyn W. Mitchell, Alex Shen, Yuko Wada, Asami Kashiwa, Lia Crotti, Federica Dagradi, Giulia Musu, Carla Spazzolini, Raquel Neves, J. Martijn Bos, John R. Giudicessi, Xavier Bledsoe, Eric R. Gamazon, Megan Lancaster, Andrew M. Glazer, Bjorn C. Knollmann, Dan M. Roden, Jochen Weile, Frederick Roth, Joe-Elie Salem, Nikki Earle, Rachael Stiles, Taylor Agee, Christopher N. Johnson, Minoru Horie, Jonathan Skinner, Michael J. Ackerman, Peter J. Schwartz, Seiko Ohno, Jamie I. Vandenberg, Brett M. Kroncke

**Affiliations:** Vanderbilt University School of Medicine, Medical Scientist Training Program, Nashville, TN, USA; Mark Cowley Lidwill Research Program in Cardiac Electrophysiology, Victor Chang Cardiac Research Institute, Darlinghurst, NSW, Australia; School of Clinical Medicine, UNSW Sydney, Darlinghurst, NSW, Australia; Department of Cardiovascular Medicine, Kyoto University Graduate School of Medicine Kyoto, Japan; IRCCS, Istituto Auxologico Italiano, Center for Cardiac Arrhythmias of Genetic Origin and Laboratory of Cardiovascular Genetics, Milano, Italy; Department of Molecular Pharmacology & Experimental Therapeutics (Windland Smith Rice Sudden Death Genomics Laboratory), Mayo Clinic, Rochester, MN, USA; Department of Physiology, University of Auckland, Auckland, New Zealand; Vanderbilt Center for Arrhythmia Research and Therapeutics, Division of Clinical Pharmacology, Department of Medicine, Vanderbilt University Medical Center, Nashville, TN, USA; Department of Medicine and Surgery, University Milano Bicocca, Milan, Italy; Division of Genetic Medicine, Department of Medicine, Vanderbilt University Medical Center, Nashville, TN, USA; Department of Molecular Genetics, University of Toronto, Toronto, ON M5S 1A8, Canada; Department of Cardiovascular Medicine, Hôpital Bichat, APHP, Université de Paris Cité, Paris, France; Department of Medicine, University of Auckland, Auckland, New Zealand; Department of Cardiology, Waikato Hospital, Hamilton, New Zealand; Department of Chemistry, Mississippi State University, Starkville, MS 39759, USA; Department of Cardiovascular Medicine, Shiga University of Medical Science, Shiga, Japan; Sydney Children’s Hospital Network, University of Sydney, Sydney, Australia; Department of Bioscience and Genetics, National Cerebral and Cardiovascular Center, Osaka, Japan

**Keywords:** LQTS, multiplexed assay of variant effect, automated patch-clamping, risk stratification, arrhythmias

## Abstract

**Background:** Long QT syndrome (LQTS) is a lethal arrhythmia syndrome, frequently caused by rare loss-of-function variants in the potassium channel encoded by *KCNH2*. Variant classification is difficult, often owing to lack of functional data. Moreover, variant-based risk stratification is also complicated by heterogenous clinical data and incomplete penetrance. Here, we sought to test whether variant-specific information, primarily from high-throughput functional assays, could improve both classification and cardiac event risk stratification in a large, harmonized cohort of *KCNH2* missense variant heterozygotes.

**Methods:** We quantified cell-surface trafficking of 18,796 variants in *KCNH2* using a Multiplexed Assay of Variant Effect (MAVE). We recorded KCNH2 current density for 533 variants by automated patch clamping (APC). We calibrated the strength of evidence of MAVE data according to ClinGen guidelines. We deeply phenotyped 1,458 patients with *KCNH2* missense variants, including QTc, cardiac event history, and mortality. We correlated variant functional data and Bayesian LQTS penetrance estimates with cohort phenotypes and assessed hazard ratios for cardiac events.

**Results:** Variant MAVE trafficking scores and APC peak tail currents were highly correlated (Spearman Rank-order ρ = 0.69). The MAVE data were found to provide up to *pathogenic very strong* evidence for severe loss-of-function variants. In the cohort, both functional assays and Bayesian LQTS penetrance estimates were significantly predictive of cardiac events when independently modeled with patient sex and adjusted QT interval (QTc); however, MAVE data became non-significant when peak-tail current and penetrance estimates were also available. The area under the ROC for 20-year event outcomes based on patient-specific sex and QTc (AUC 0.80 [0.76-0.83]) was improved with prospectively available penetrance scores conditioned on MAVE (AUC 0.86 [0.83-0.89]) or attainable APC peak tail current data (AUC 0.84 [0.81-0.88]).

**Conclusion:** High throughput *KCNH2* variant MAVE data meaningfully contribute to variant classification at scale while LQTS penetrance estimates and APC peak tail current measurements meaningfully contribute to risk stratification of cardiac events in patients with heterozygous *KCNH2* missense variants.

**Clinical Perspective:** *What is new?:*

- A two-order of magnitude increase in the set of calibrated functional data for *KCNH2*-LQTS is provided by two complementary *KCNH2* assays
- Proactively available variant scores are presented for all possible missense variants by using a LQTS penetrance estimation framework conditioned on high-throughput MAVE data
- Variant functional data, in addition to patient features of corrected QT interval and sex, significantly improve modeling of 20-year cardiac event outcomes

*What are the clinical implications?:*

- Readily available MAVE scores for thousands of variants may facilitate classification of new variants discovered in individuals with suspected LQTS
- Scores and penetrance estimates are readily searchable at variantbrowser.org for community inquiry
- Both automated patch-clamp data and quantitative LQTS penetrance estimates, conditioned on MAVE data, improve prediction of 20-year cardiac event outcomes in a large cohort of *KCNH2* heterozygotes

## Introduction

Long QT syndrome (LQTS) is a lethal arrhythmia condition, with nearly 30% of cases caused by rare loss-of-function variants in the cardiac potassium channel gene *KCNH2*^1,2^. Variant classification is difficult, often owing to lack of functional data. Risk stratification for patients heterozygous for these variants is also complicated by heterogenous clinical data and incomplete penetrance. Variant-specific functional data can assist with variant classification and diagnosis^3,4^; however, data are not available at a scale commensurate with their discovery in affected and unaffected populations. In addition, whether variant-specific information associates with disease-relevant presentations, including adverse outcomes, is unknown. These challenges present barriers to effective genotype-first medicine^5,6^, where large sequencing efforts increasingly identify individuals heterozygous for previously unrecognized variants in clinically actionable genes, including *KCNH2*^7^. By providing more granular variant-specific data, a clinician could ideally improve diagnosis and cardiac event risk stratification when also considering classic patient-specific features such as corrected QT interval and patient sex, as in the case of LQTS.

Most *KCNH2* loss-of-function variants disrupt membrane trafficking but may also cause gating defects and other changes in ion permeability^8^. Here, we deployed two high-throughput functional assays to assess *KCNH2* variant effect (Figure 1): a proactive Multiplexed Assay of Variant Effect (MAVE) to quantify channel concentration at the plasma membrane (18,796 variants), and detailed Automated Patch-clamping (APC) to record potassium current densities (from 533 variants previously identified in LQTS patients). We demonstrated the utility of these MAVE data in *KCNH2* missense variant classification, evaluated against a curated set of clinically-ascertained variants and ClinVar^9^. Beyond classification, we tested the hypothesis that variant-specific data, including these high-throughput functional data and our previously described Bayesian LQTS penetrance estimates^10-12^, associate with patient cardiac events. Accordingly, we curated an international cohort of 1,458 *KCNH2* missense variant heterozygotes with detailed cardiac event history from arrhythmia centers (ascertained) and the UK Biobank (unascertained). We find that time-to-event models including these variant-specific data significantly associated with cardiac events, when controlling for traditionally used patient-specific features of corrected QT interval and sex. These proactive data are now publicly available (variantbrowser.org) to support variant classification and event stratification for *KCNH2* variant heterozygotes.

**Figure 1.**
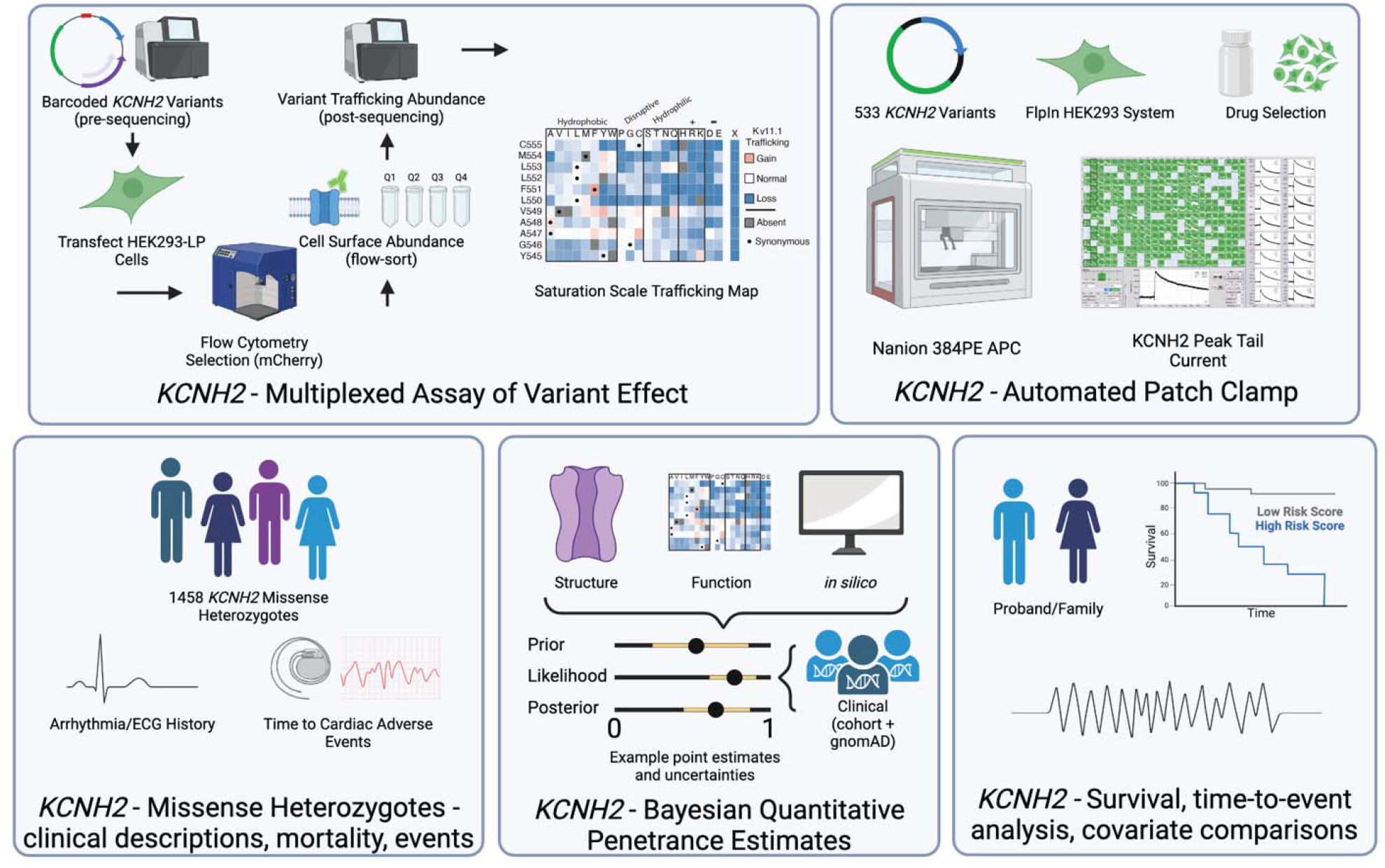
Project Overview. Integration of two high-throughput functional assays with clinical deep phenotyping, quantitative penetrance estimates, and prospective risk prediction models.

## Methods

Full methods are described in the online Supplemental Methods and Appendix. All clinical data used in this analysis received IRB/REC approval. The research reported in this paper adhered to guidelines included in the Helsinki Declaration as revised in 2013. All code used to analyze these data may be found on GitHub at https://github.com/kroncke-lab.

## Results

### Full Implementation of a Multiplexed Assay of Variant Effects to Quantitate Trafficking

We previously designed and piloted^13,14^ a high-throughput MAVE to quantitate the plasma membrane concentration of *KCNH2* variants in HEK cells. Here, we characterized 18,796 variants^13-15^ and found excellent stratification of WT-normalized variant MAVE scores across synonymous (mean 100.1±24.2%), missense (mean 71.7±46.6%), and nonsense (mean 17.9±36.0%) variant classes (Figure 2A-C; Supplemental Tables 1-2). Scores were heterogenous throughout the protein, even within ‘hot-spot’ domains. Nonsense variants invariably showed almost complete loss of function up to residue 863 (1.35 ± 7.86%)) but minimal impact on protein trafficking distal to residue 863 (71.7 ± 39.1%; Supplemental Figure 1)^8^.

**Figure 2.**
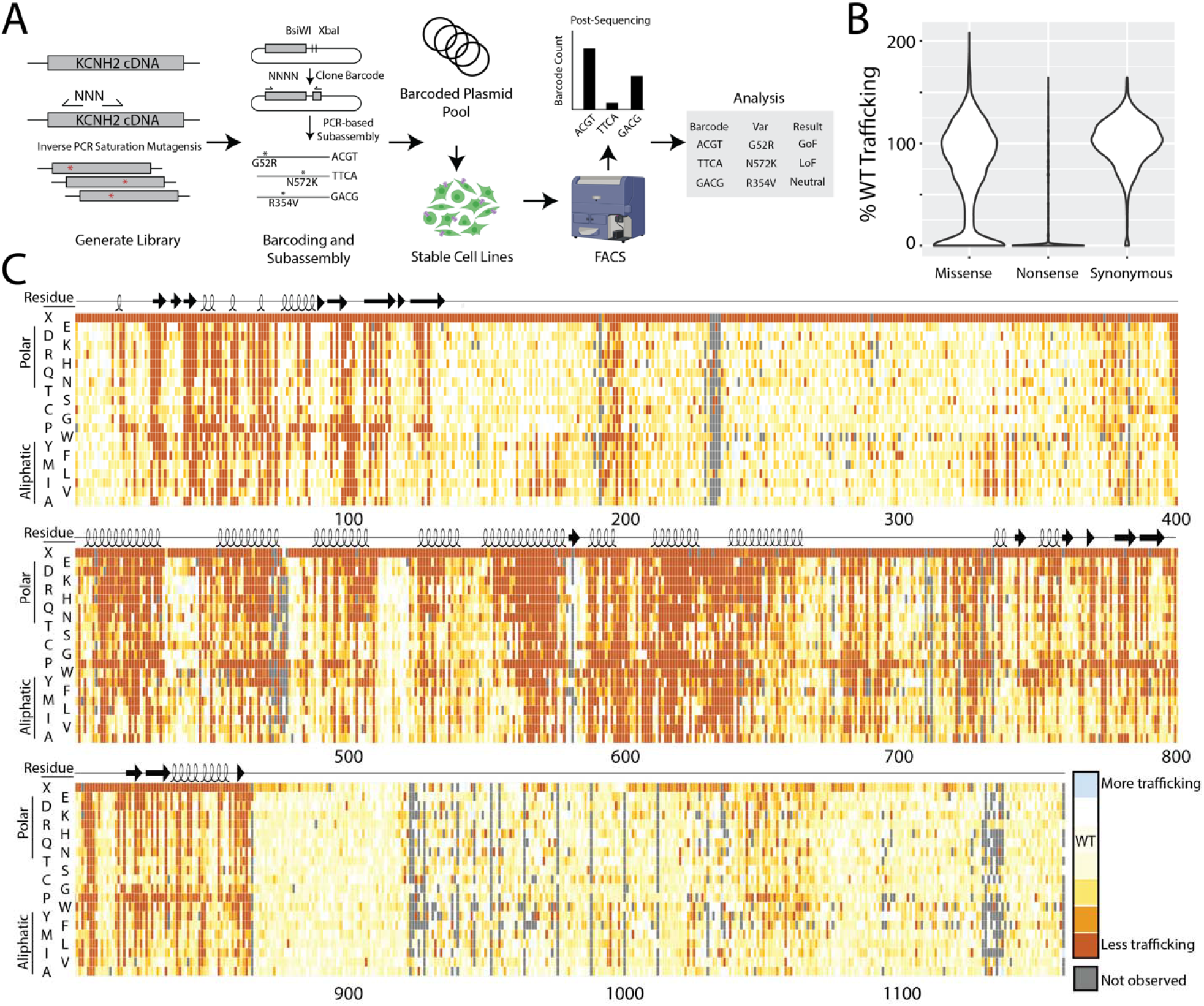
Results of a *KCNH2* Multiplexed Assay of Variant Effect. **A)** Schematic of MAVE assay. We employed a barcode abundance-based MAVE of cell-surface *KCNH2* variant expression to quantify variant trafficking, the primary mechanism of *KCNH2* variant loss-of-function. **B)** Distribution of WT-normalized variant trafficking scores among missense, synonymous, and nonsense variants. **C)** Heatmap depicting trafficking scores across the coding region of *KCNH2*. Dark orange indicated less than WT trafficking, white similar trafficking to WT, and blue increased trafficking. Missing data are depicted in gray.

To determine the appropriate application strength for benign or pathogenic variant functional evidence, we performed calibration of the MAVE data. First, we calibrated to ClinGen-recommended guidelines using Odds of Pathogenicity (OddsPath) as previously reported^16^. We used the same curated controls for MAVE OddsPath calibration as those for our previously published APC calibration; abnormal/normal were determined by z-score thresholds from the mean distribution of scores among Benign/Likely Benign control variants (Figure 3A and 3B)^16,17^. A z-score range of -2 to 2 corresponded to trafficking scores of 58% to 153% of WT. Using these thresholds as normal function, we found excellent MAVE stratification of B/LB and P/LP controls, yielding up to strong pathogenic functional evidence (OddsPath 21.3) and up to moderate benign functional evidence (OddsPath benign 0.122; Figure 3A and 3B; Supplemental Table 3). Next, we mapped log likelihood ratios to ACMG evidence codes using the approach of Tavtigian et al.^18^ (Supplemental Figure 3 and Supplemental Table 4). This second approach would indicate application of ‘very strong’ pathogenic functional evidence for variants with trafficking scores less than 35% of WT scores. Beyond the curated list of controls, our MAVE assay had high functional evidence strength across all Benign/Likely Benign and Pathogenic/Likely Pathogenic variants within ClinVar (AUC 0.977 [0.957, 0.998]; N=152; Figure 3C).

**Figure 3.**
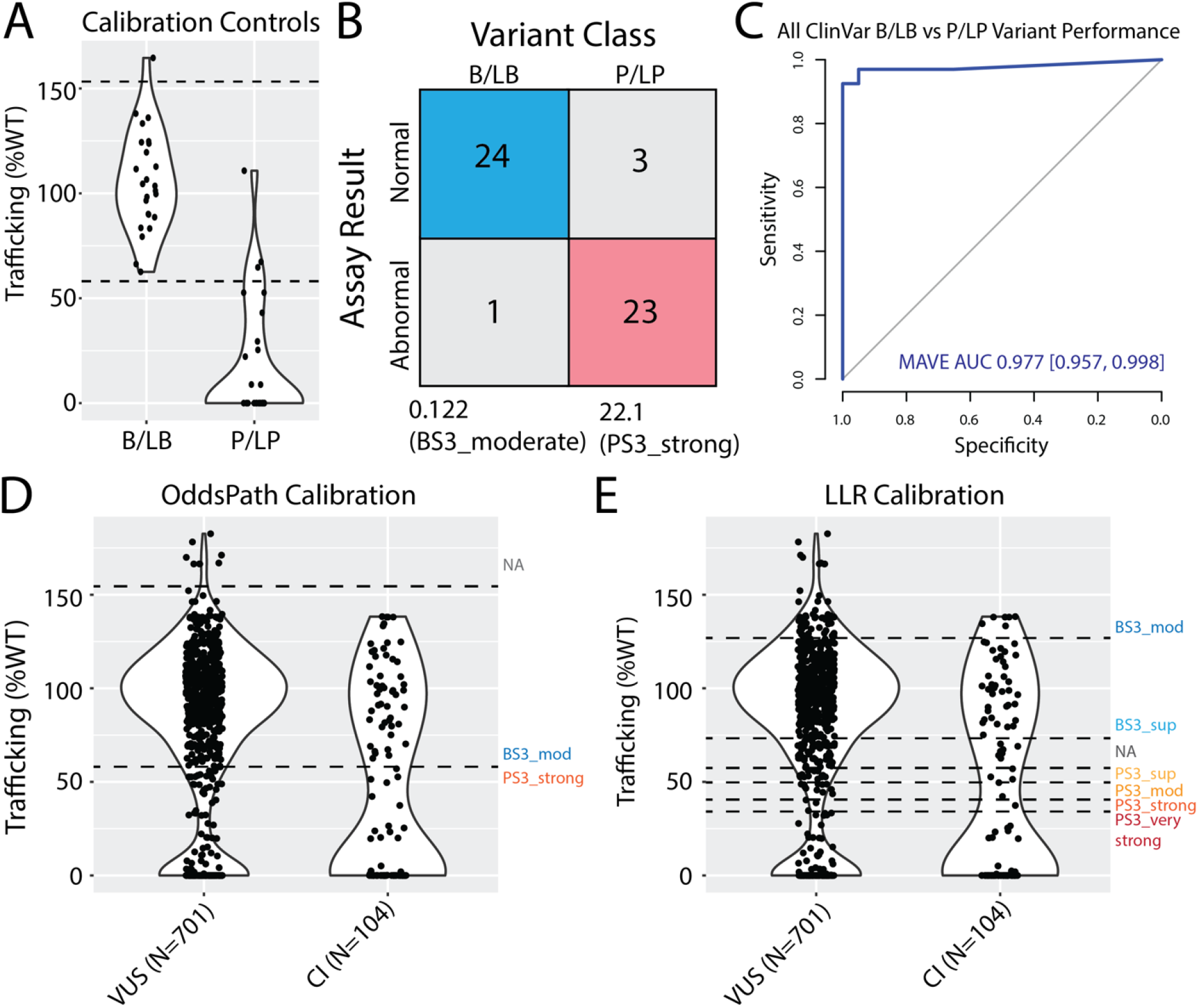
Classification and Diagnostic Value of MAVE Trafficking Assay. **A)** Distribution of trafficking scores among control variants for assay calibration. Controls selected from those described by Thomson et al. **B)** 2×2 table of assay result vs variant classification in control group. An abnormal assay result was defined as z-score < -2, arising from the distribution of B/LB variant scores as previously described. **C)** Receiver operator characteristic curve of MAVE data applied across all readily available ClinVar B/LB and P/LP annotations. **D-E)** Violin plots showing trafficking scores of VUS and Conflicting Interpretation variants in ClinVar. Dotted lines reflect OddsPath thresholds **(D)**, and Log-likelihood ratios **(E)**.

Next, we tested the prospective value for reducing clinical uncertainty among variants in ClinVar using both calibrations. First, we found that 160 of 701 missense Variants of Uncertain Significance in ClinVar had strong pathogenic functional scores by OddsPath, and 130 of 701 by LLR (*pathogenic strong*, Figure 3E). Critically, 415 and 55 of 701 would achieve supporting and moderate benign functional evidence, respectively. An increasingly complicated class of variants are those with Conflicting Interpretations in ClinVar, for which functional data can serve as an effective arbiter. A total of 49 of 104 Conflicting Interpretation variants meet strong pathogenic functional evidence criteria, whereas 37 and 6 meet benign supporting and benign moderate criteria. Together, these data significantly address the current classification problem.

### Automated Patch-Clamping to study KCNH2 Variant Peak Tail Currents

To measure variant effects beyond trafficking, we used the SyncroPatch 384 PE platform^17,19^. We comprehensively interrogated potassium peak tail currents among *KCNH2* variants observed in our clinical cohorts supplemented with some common variants found in gnomAD^20^ (Figure 4A; Supplemental Methods). 331/533 variants had loss-of-function (z < -2, Figure 4B, Supplemental Table 5). We observed good concordance of variant effect, as determined by independently derived z-scores, between MAVE and APC datasets, including across “hot spot” domains (Figure 4C-D; Supplemental Figure 4). 388 of 443 (90%) variants studied in both assays were concordantly annotated with a normal or abnormal threshold of an assay Z-score less than -2. Twenty three of 443 had normal trafficking but abnormal current density, suggesting abnormalities in gating or ion permeability.

**Figure 4.**
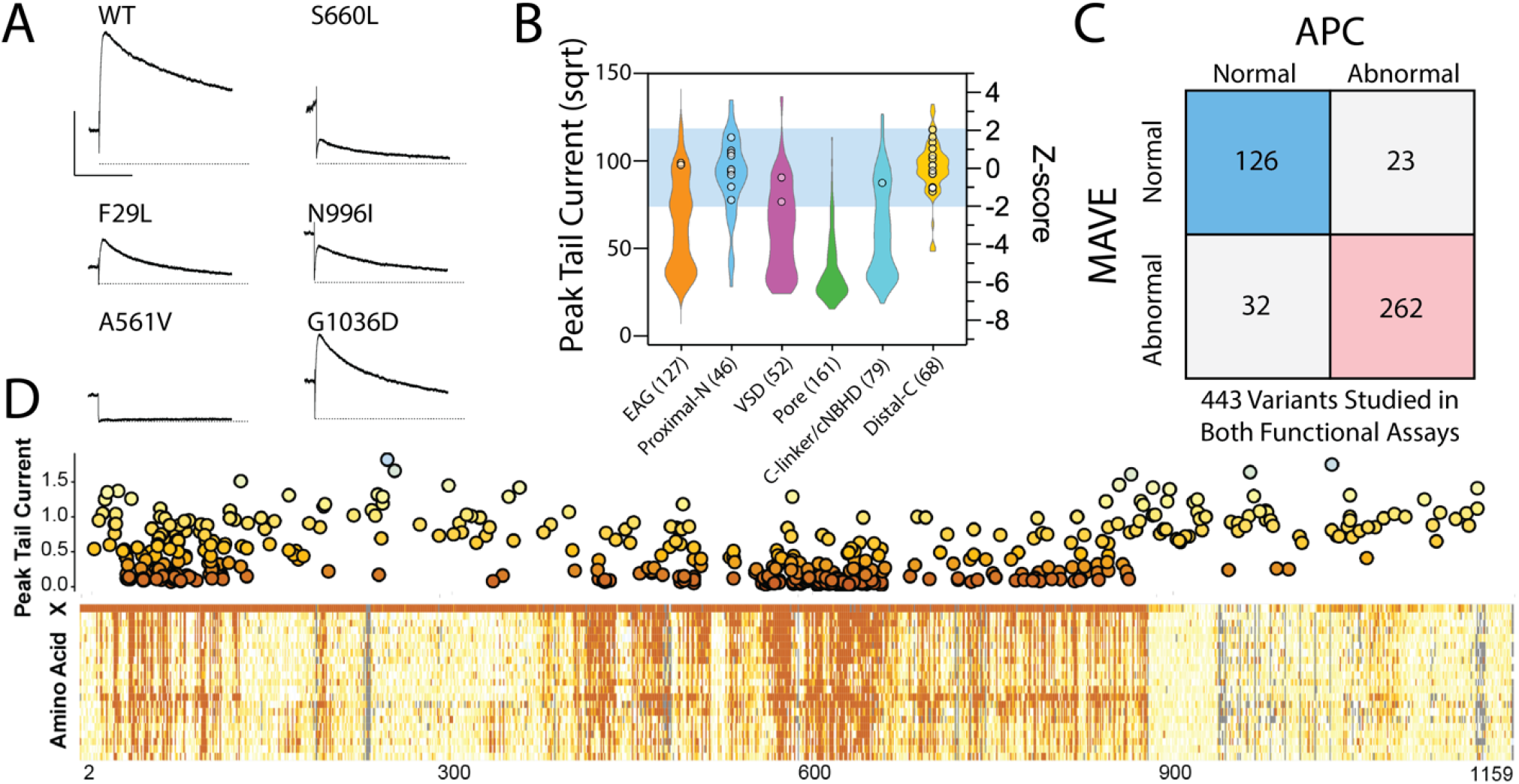
Results of a *KCNH2* Automated Patch Clamp assay. **A)** Example APC peak tail currents recorded at -50 mV showing different levels of function. Y-axis is 500 pA and X-axis is 500ms. **B)** KCNH2 peak-tail current densities for 533 variants (n=38,772 recordings) across 6 domains of the protein observed in our clinical cohort, gnomAD^20^, and previous literature reports^27^. Benign variant controls from gnomAD are shown as white circles. Blue range depicts variants with ‘normal function’, as defined by a ±2 z-score window from the mean current density for B/LB variants^17^. **C)** Matrix of z-score determined normal and abnormal variants studied by both functional assays. **D)** Visual correlation of functional assays by residue position.

### LQTS Penetrance Estimates

Our group has previously described a Bayesian approach to estimate LQTS variant penetrance, the estimated fraction of affected over total individuals harboring a variant^10,11^. This approach first leveraged low-throughput functional data, *in silico* predictions, and channel structural features^12^. We now condition the LQTS penetrance prior estimates on MAVE functional data to further improve these predictions and leverage *all* proactively available variant-specific data and clinically phenotyped individuals in the literature (see Supplement and references 10-12 for detailed descriptions). Notably, penetrance estimates are now available for all possible *KCNH2* missense variants. We describe these estimates as ‘LQTS Penetrance’ throughout the text.

### Correlation of Functional Data with LQT2 Clinical Endophenotypes

To test the association of these MAVE and APC data with relevant clinical features, we curated a large, deeply phenotyped cohort of 1,458 patients recruited from four tertiary arrhythmia clinics in Japan (N=289), Italy (N=275), the USA (N=261), and New Zealand (N=66) and the UK Biobank (N=574) heterozygous for 418 unique *KCNH2* missense or in-frame insertion/deletion variants (Table 1). At baseline, cohort QTc values followed expected trends, with higher mean values among probands, females, and individuals experiencing cardiac events (Supplemental Figure 5; events defined as syncope, ventricular tachycardia, ventricular arrhythmias, ICD shocks). MAVE scores (Spearman’s ρ 0.59 [0.54, 0.63]), APC measured peak tail current (Spearman’s ρ 0.61 [0.57, 0.65]), and LQTS penetrance estimate (Spearman’s ρ 0.68 [0.65, 0.71]) each significantly correlated with corrected QT interval (Figure 5A-C). We found that individuals experiencing cardiac events had statistically significantly more deleterious mean MAVE trafficking, LQTS penetrance estimates, and APC peak current densities (Figures 5D-F).

**Table 1.** Demographics and clinical features of cohort for individuals heterozygous for variants with available MAVE and APC data. Standard deviations presented in parentheses. N indicates total amount of heterozygotes across cohort for each category.

|  |  | QTc (ms) | MAVE | APC | N |
| --- | --- | --- | --- | --- | --- |
| Event |  |  |  |  |  |
|  | No | 458 (64.5) | 54.3 (45.9) | 58.6 (37.4) | 1204 |
|  | Yes | 516.8 (62.2) | 26.7 (39.8) | 27.5 (31.5) | 254 |
| Sex |  |  |  |  |  |
|  | Female | 478 (54.7) | 45.9 (45.8) | 49.2 (37.9) | 746 |
|  | Male | 462.2 (80.1) | 51.5 (46.4) | 55.2 (38.6) | 712 |
| Country |  |  |  |  |  |
|  | Italy | 476.6 (43.7) | 42.4 (44.3) | 51.5 (41.7) | 268 |
|  | Japan | 492.8 (106.7) | 34.8 (44.9) | 38.6 (38.4) | 289 |
|  | NZ | 508.7 (67.2) | 33.2 (44.4) | 37.8 (38.2) | 66 |
|  | UKBB | 425.5 (24.6) | 86.5 (33.8) | 79.8 (22.6) | 574 |
|  | USA | 489.5 (45.9) | 24.5 (32.5) | 34.5 (30.6) | 261 |

**Figure 5.**
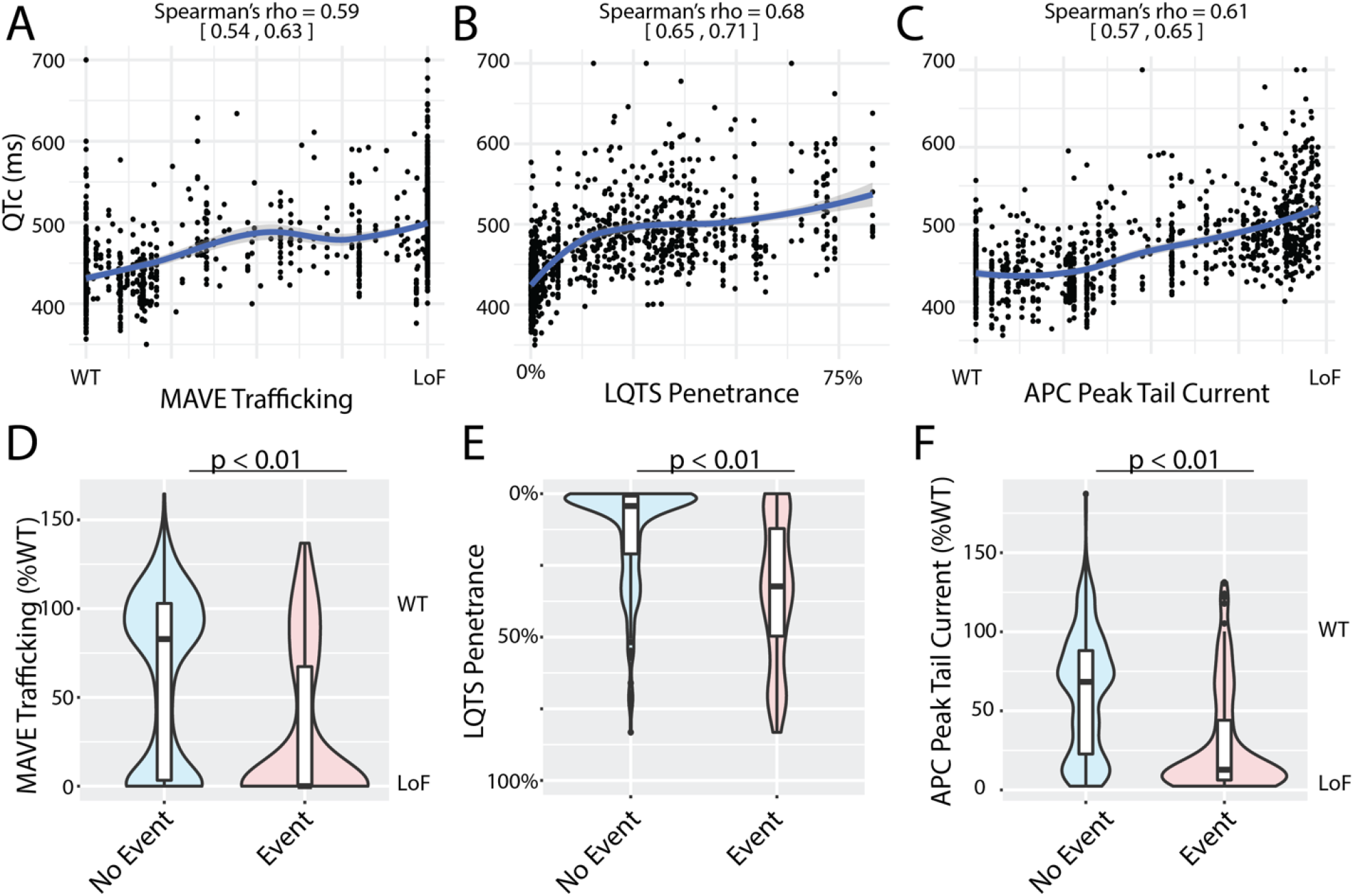
Descriptive Correlations of Functional Data with Missense Heterozygote Cohort Clinical Features. **A-C)** Correlations between participant QTc and functional scores for all available cohort members with each descriptor (Spearman rho). **D-F)** Stratification of each descriptor by cohort members experiencing cardiac event (Mann-Whitney U-test).

### Predictive Application of Functional Data to Cardiac Event Risk Stratification

We next sought to test the utility of these variant-specific features in cardiac event risk stratification when combined with clinically available patient data of QTc and sex. In a multivariate Royston-Parmar time-to-event model including corrected QT interval and sex, the newly described MAVE scores, APC measured peak tail current, and LQTS penetrance estimate (Figure 6A-C) each independently associated with LQTS-related cardiac events. However, MAVE scores were insignificant when all three variant-specific features were combined as a single model (Figure 6D), as was observed by variant-specific covariate selection using Least Absolute Shrinkage and Selection Operator (Supplemental Methods and Supplemental Figures 6-7). The area under the ROC for time-to-event model (evaluated at 20 years) of patient-specific sex and QTc (AUC 0.80 [0.76-0.83]) was improved with LQTS penetrance estimates (AUC 0.86 [0.83-0.89]) or attainable peak tail current data (AUC 0.84 [0.81-0.88]; Figure 6E); Harrel’s c-statistic, Log-likelihood, and Akaike information criterion were also significantly improved by addition of LQTS penetrance estimates or attainable peak tail current data (Supplemental Table 6).

**Figure 6.**
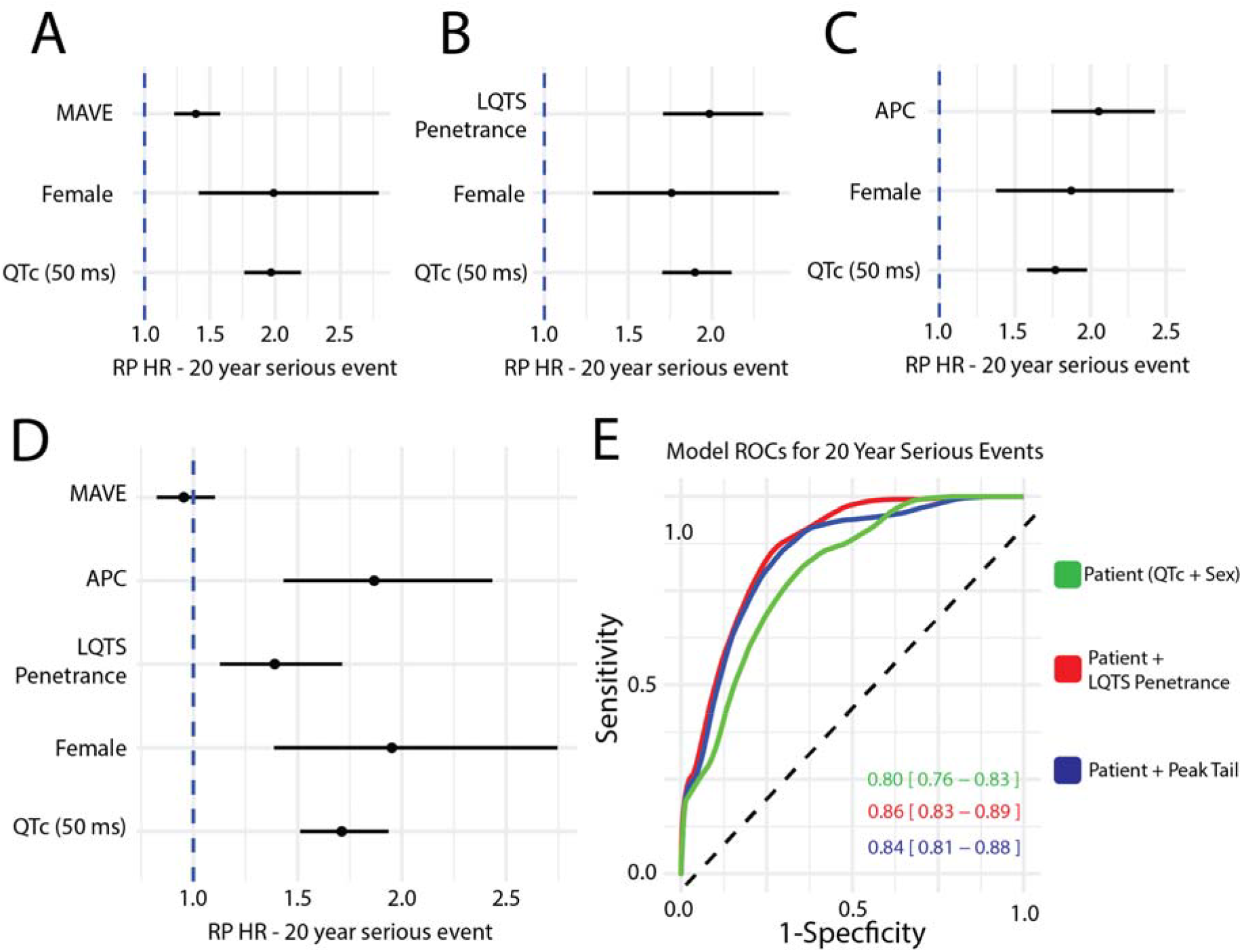
Clinical Risk Models and Applications of Variant-specific and Patient-Specific features. **A-C)** Royston-Parmar Hazard Ratios (RP HR) for first 20-year cardiac event with baseline patient-specific features of sex and adjusted QTc, and variant-specific data of MAVE, LQTS penetrance, and APC. **D)** Royston-Parmar Hazard Ratios (RP HR) for first 20-year cardiac event cardiac event with all patient-specific and variant-specific data. **E)** ROCs/AUCs for three different models for all cardiac events through 20 years of age.

## Discussion

We present a comprehensive functional analysis of 18,796 missense variants in a ClinGen definitive evidence LQTS-associated gene^2^, increasing the available set of calibrated functional data for *KCNH2*-LQTS variants by two orders of magnitude. The functional data and penetrance estimates are readily searchable at variantbrowser.org for community inquiry which will have an immediate clinical impact in assisting with reclassification of variants of uncertain significance. We also demonstrate that integrating continuous and quantitative variant-specific features with clinical data improves modeling of 20-year cardiac event outcomes.

This work uniquely complements previous MAVE projects in cardiac disease to classify variants^21-23^, and lower throughput manual patch-clamp data to risk stratify patients^24^. For example, previous MAVE data has been used to study variants in the LQTS-associated^2^ genes *KCNE1*^21^ and *CALM1-3*^23^. An interesting clinical application of the *CALM1-3* MAVE was the readily available map to facilitate interpretation of VUS later observed in patients undergoing genetic evaluation for cardiac arrest^25^. In an impressive study, Ishikawa et al. used manual patch-clamp to functionally study and classify 55 *SCN5A*-Brugada Syndrome variants, and then risk stratify affected Japanese heterozygotes for ventricular arryhtmias^24^. The current study greatly expands on the number of variants evaluated for classification, and then applies these data for event risk stratification in a larger, international LQTS cohort. Compared to previous works, the current study also emphasizes the value of complementary assays. For example, we observe that MAVE data performs near equally to APC for variant classification; however, based on functional data alone, APC has higher performance for event risk stratification.

Our findings directly complement the actionability of recently described European Society of Cardiology guidelines for management of ventricular arrhythmias and sudden cardiac death^26^. For example, the improved classification of variants enabled by these high-throughput studies will facilitate the Class I, Level C recommendation of LQTS diagnosis in the presence of any pathogenic variant. Furthermore, this same decrease in clinical uncertainty through classification may improve a Class IIa, Level B recommendation of beta-blocker treatment for patients with a pathogenic variant and normal QTc. Lastly, a Class IIa, Level C recommendation for risk stratification and therapy initiation may be improved from ‘genotype’ information, to more granular, variant-specific information.

### Limitations

Experimental work was performed in HEK293 cells which do not fully reflect variant effect in native cardiomyocytes. Nevertheless, both high throughput functional assays were able to accurately distinguish between known pathogenic and known benign variants. Heterozygotes from arrhythmia cohorts suffer from ascertainment bias and may have experienced more events than other individuals with identical variants (incomplete penetrance). Ideally, the utility of these assays should be prospectively studied in large scale cohorts of genotype-first guided medicine.

## Conclusion

High-throughput variant functional data improves diagnosis/variant classification, while APC peak tail current and LQTS penetrance estimates improve modeling of cardiac event outcomes for *KCNH2*-LQTS.

## Supporting information

Supplement Doc

## Data Availability

All non-participant-level data are contained in the manuscript. Data analysis pipelines are fully available at https://github.com/kroncke-lab. Prospective data for KCNH2 variant interpretation are hosted at https://variantbrowser.org/.

https://github.com/kroncke-lab

## Data and Code Availability

Code used to analyze raw data and generate figures and tables are available at the Kroncke Lab GitHub site. Bayesian variant functional scores for MAVE and APC, *in silico* predictions, structural features, and clinical data are available at variantbrowser.org.

## Acknowledgements

We thank Kenneth Matreyek and Doug Fowler for the HEK293 landing pad cells. Figure 1 was made using BioRender.

