## Supplement Doc for "Multiplexed Assays of Variant Effect and Automated Patch-clamping Improve *KCNH2*-LQTS Variant Classification and Cardiac Event Risk Stratification"

**Supplemental Methods**

Multiplexed Assay of Variant Effect – Quantification of *KCNH2* Variant Trafficking

Automated Patch Clamp – *KCNH2* Variant Peak Tail Current Measurements

Clinical Analyses – Phenotyping, Demographics, Survival, and Hazard Ratios

**Supplemental Discussion**

**Supplemental Figure 1.** Trafficking Scores of Nonsense Residues Before and After Residue 863.

**Supplemental Figure 2.** Structural Heterogeneity of “hot spot” Domains.

**Supplemental Figure 3.** Log-likelihood ratios derivation for MAVE.

**Supplemental Figure 4.** Correlation of MAVE and APC Functional Scores.

**Supplemental Figure 5.** Plots of cohort corrected QTc intervals.

**Supplemental Figure 6.** Co-linearity analysis of full model covariates by Spearman correlation analysis.

**Supplemental Figure 7.** LASSO regularization path analysis.

**Supplemental Table 1.** Sequences of Primers and Plasmids used in this study.

**Supplemental Table 2:** MAVE Trafficking Scores.

**Supplemental Table 3:** OddsPath for MAVE assay.

**Supplemental Table 4:** Log-likelihood ratios for MAVE assay.

**Supplemental Table 5:** APC peak tail current measurements.

**Supplemental Table 6:** Information criteria for risk stratification models.

**MAVE Experiments**

*Library Saturation Mutagenesis.* We conducted a comprehensive mutagenesis of the *KCNH2* gene, employing previously standardized protocols^1,2^. We created a tile-based system by introducing multiple restriction sites into the coding region of *KCNH2* using the Quickchange Lightning Multi kit (Agilent). The starting segment was a wild-type *KCNH2* plasmid in AttB-KCNH2-HA:IRES:mCherry^2^. To enable cell surface labeling of K_V_11.1, we inserted an HA tag (NSEHYPYDVPDYAVTFE) between amino acids T443 and E444. We performed saturation mutagenesis with mutagenic primer pairs. These were designed with a 5'NNN segment on the forward primer, where N represented an equal mix of A/C/G/T. The primary objective was to generate a diverse pool of plasmids containing a balanced assortment of possible codon combinations. To achieve this, we obtained a set of all the mutagenesis primers covering all the amino acids of K_V_11.1, and after the mutagenesis, we pooled the resulting products. Following PCR purification, the products were subjected to phosphorylation using T4 Polynucleotide Kinase (New England Biolabs, NEB) at 37ºC for 1 hour. Subsequently, we carried out ligation with T4 DNA ligase (NEB) for 16 hours at 16ºC, followed by digestion with DpnI (NEB) for 1 hour at 37ºC. The mutagenized PCR products were then purified and electroporated into MegaX DH10B Electrocomp cells (ThermoFisher) using a gene pulser electroporation system (Mod. No. 1652076, BioRad). The electroporation conditions were set at 25uF, 200 ohms, and a pulse of 2V for approximately 5 seconds. To assess the success of the procedure and quantify the quality and diversity of plasmid uptake, we grew colonies after electroporation and conducted parallel colony expansion in 50 mL of broth. The colonies were cultivated on LB-ampicillin agar plates at a dilution of 1:10,000, and to further analyze the diversity of the mutant library for each experiment, we performed Sanger sequencing on 10 selected colonies. To incorporate the mutant tile libraries into the full *KCNH2*, we carried out subcloning of the tile into the AttB-KCNH2-HA:IRES:mCherry plasmid through restricted digestions. The digested product was ligated with T4 ligase (NEB) and introduced into cells using the standard electroporation protocol.

*Barcode Generation and Linking to KCNH2 Variants.* We used subassembly to link barcodes to each *KCNH2* variant within the pool. Using i7 and i5 primer pairs with specific adapter sequences, we performed PCR to generate multiple pools of sequences containing the barcodes and variant sequences in close proximity. PCR products were purified using Ampure XP beads (Beckman Coulter) according to the manufacturer's protocol. The purified PCR products were sequenced on an Illumina NovaSeq sequencer at the VANTAGE facility, VUMC. The resulting data were obtained in fastq format and analyzed using inhouse Python and R scripts (https://github.com/kronckelab/KCNH2_DMS).

*Transfecting and Creating Stable Cell Lines.* We generated stable cell lines incorporating a single copy of *KCNH2* per cell using ‘landing pad’ HEK293T cells^3,4^ (kind gift of Kenneth Matreyek and Douglas Fowler). Cells were cultured in alpha MEM (Corning), enriched with 10% FBS (Gibco) and 1% GlutaMax (Gibco), and maintained at a constant temperature of 37 ºC with 5% CO_2_. Cells were grown to a confluency of 40% to 50% prior to transfection according to manufacturer’s instructions with FuGene 6 (Promega) in OptiMEM (Gibco). Two plasmids were co-transfected: one expressing Bxb1 integrase (pCAG–NLS–HA–Bxb1; Addgene #51271, generously provided by Pawel Pelczar) and another containing the barcoded pool of *KCNH2* variants.

*Preparation of Cells for Flow Sorting-Based Trafficking Analysis.* To induce K_V_11.1 expression, cells were treated with 1µg/ml doxycycline for 24 hours before flow sorting. The doxycycline-induced HEK293T-integrated library was sorted using fluorescence-activated cell sorting (FACS) to obtain cells expressing mCherry, indicating successful integration of the AttB-containing plasmid. Cells were dissociated from the 6-well-plate (Corning) using TrypLE Express (Thermo Fisher), resuspended in alpha MEM, and centrifuged at 200 g for 5 minutes. The pellet was resuspended in ~1 mL of phosphate-buffered saline (PBS) containing 1% Bovine Serum Albumin (BSA) and filtered using a 5 ml polystyrene round-bottom tube with a cell strainer cap (Corning). The cell population was quantified using an automated cell counter (Biorad), and diluted to a concentration of 3-4 million cells per 1 mL with 1% BSA PBS solution. These cells were incubated in mouse anti-HA antibody conjugated to Alexa 647 (Cell Signaling Technology) at a dilution of 1:500, shaken for 15-30 minutes, and pelleted at 200 g for 5 minutes. Cells were then resuspended in 1% BSA PBS, filtered again, and FACS sorted for mCherry positive and BFP negative cells, confirming successful plasmid integration. The cells were collected as 4 separate aliquots based on the intensity of Alexa 647, denoted as low expression (L), medium expression (M), high expression (H), and expression-negative (Neg) cells. The desired cells were sorted into 5 ml collection tubes, transferred to 6-well plates (Cellstar), and treated with 1x penicillin-streptomycin solution (Gibco). The cells were grown for 4 days until reaching a confluency of approximately 2 million cells per well. They were then dissociated with TrypLE Express (Gibco), pelleted at 200 g for 5 minutes, resuspended in PBS, and cryopreserved at approximately 1 million cells per mL, per vial.

*DNA Isolation and Library Preparation for Illumina Sequencing.* DNA was isolated from each of the H, M, L, and Neg sorted pools using 100 μL QuickExtract (Lucigen) per 1 million cells, following the manufacturer’s instructions. PCR, with 25-35 cycles, was carried out using 2.5 μL of isolated DNA and Q5 polymerase (NEB), according to the manufacturer’s instructions. The resulting DNA libraries were purified with AmpureXP beads following the manufacturer’s instructions. The libraries were sequenced on a NovaSeq 6000 instrument with 150 base paired-end sequencing.

*Biological and Technical Replication.* For each tile, we created two independent barcoded *KCNH2* variant libraries with unique barcode-variant associations. These plasmid pools, were subsequently transfected into three independent HEK293 cell lines, resulting in a total of 6 biological replicates. Within each plasmid, we observed a range of approximately 2 to 9 technical replicates (multiple barcodes per variant), representing the interquartile range. The redundancy of barcodes and codon substitutions linked to the same missense variant accounted for this variation in the number of technical replicates. This approach allowed us to capture inherent variability and ensure comprehensive coverage across underrepresented codons.

*Assigning Trafficking Scores.* Variant counts from each pool of sorted cells were aggregated to calculate a trafficking score using the following equation:

$${Trafficking score}_{i}=\frac{1*\left( {Pool}_{1,i} \right)+2*\left( Pool_{2,i} \right)+3*\left( Pool_{3,i} \right)+4*(Pool_{4,i})}{{Total number of barcodes observed}_{i}}$$

Where Pool_n,i_ is the fraction of the ith barcode in the nth pool of sorted cells; n ranges from 1 (no K_V_11.1 present, i.e. no AF647 signal) to 4 (high abundance of KV11.1, i.e. high AF647 signal). After obtaining the individual scores, we proceeded to aggregate them by variant and subjected them to a normalization process through a linear transformation. This ensured that the trafficking scores were scaled to a range of 0 (representing barcodes exclusively observed in the AF647 negative pool) to 100, corresponding to the wild-type (WT) condition (see python script at GitHub for full details). Furthermore, we averaged the scores across the two replicate experiments, which entailed separate transfections of unique barcoded *KCNH2* variant libraries. Variants with a standard error greater than 0.2 were removed from subsequent analyses.

*Heterogeneity of “Hot spot” Domains.* The PM1 ACMG criteria has been used to assign domain or “hot spot” weight for variant classification^5^. To investigate functional scores among variants with near saturation-scale evidence, we quantified variant effect using the following residues of “hot spots”: EAG: 1-135; Proximal-N: 136-400; VSD: 401-550; Pore: 551-670; C-linker/cNBHD: 671-870; Distal-C: 871-1159. We plotted functional scores using the ggplot2 package in R, and mapped onto the Cryo-EM structure of K_V_11.1 as previously described^6^.

*Z-score Calibration and determination of OddsPath.* The ClinGen Sequence Variant Interpretation working group recommended calibrating assay functional evidence strength to performance on pathogenic and benign control variants^7^. A limitation of this approach is the number of ClinVar-annotated B/LB and P/LP variants available for calibration^8^. To harmonize MAVE and APC functional assays, we used the same set of B/LB variants and P/LP variants from a recent collaboration of the K_V_11.1 APC assay^9^. We derived the OddsPath as described in the equations in Supplemental Table 4. Determination of the normal/abnormal threshold was made using the mean trafficking scores among B/LB variants in the assays and a 2 Z-score threshold in either direction^10^. These thresholds corresponded to 59-156% of WT trafficking in the MAVE assay.

*Log Likelihood Ratio*. We employed kernel density estimation (KDE) with cosine kernels and adhered to Silverman's rule of thumb for bandwidth selection to construct probability density functions of scores for positive (pathogenic) and negative (benign) reference variant sets, as described above. To mitigate the risk of erratic fluctuations in the log likelihood ratio (LLR) in score ranges where both densities approach zero, we conservatively adjusted the LLR to zero, ensuring stability in areas lacking sufficient data. Furthermore, we observed a transition in evidence codes from BS (Benign Strong) to BP (Benign Supporting) as the trafficking score exceeded 200%, reflecting the paucity of reference variants in this range and the consequent tapering of the π_benign density.

**Automated Patch Clamp Experiments**

*KCNH2* *variant plasmid DNA.* The *KCNH2* variant plasmids were designed according to a previously published protocol^11^. The generation of the *KCNH2* variant, restriction digest, subcloning into the pcDNA5/FRT/TO vector (Thermofisher) and confirmatory Sanger sequencing were completed by GenScript Inc (Pistcataway, NJ, USA). Each of the plasmid contains a copy of a unique *KCNH2* variant and a copy of WT separated by an IRES element to model the heterozygous cell state.

*KCNH2* *Flp-In HEK293 variant cell lines*. The generation of inducible Flp-In *KCNH2* variant HEK293 cell lines were performed according to a previously published protocol^11^. The *KCNH2* plasmid DNA was transfected with POG44 Flp-Recombinase Expression Vector (Thermofisher) into Flp-In HEK293 parental cell (Thermofisher) using lipofectamine 3000 (Thermofisher). The stable Flp-In *KCNH2* variant cell lines were then selected using 200ug/mL hygromycin (Thermofisher) and cryopreserved for the APC assay.

*APC protocols and data analysis*. The Nanion SyncroPatch 384 PE APC platform was used to quantify the function of *KCNH2* variants. KCNH2-expressing cells were depolarized to +40mV for 1s followed by repolarization to -50mV for 3s to record the peak tail current. The measured peak tail current represents the rapid delayed-rectifier current (*I*_Kr_) available for repolarization, that will be modified by any changes in trafficking as well as changes in the voltage-dependence and kinetics of channel activation and inactivation^12^. Channel deactivation was also captured during the 3s repolarization at -50mV. The different level of peak tail current was normalized by the respective capacitance and transformed using a square root function to achieve normality^10^. The square-root transformed data was normalized to the WT on the same assay plate to control for plate-to-plate variability.

**LQTS Penetrance Estimates**

We estimate Long QT Syndrome (LQTS) variant-specific penetrance as a continuous, quantitative probability of disease manifestation for missense variants, not binary classifications of 'pathogenic' or 'benign’ or their likelihood, as previously described^6,13,14^. Within a Bayesian framework, we calibrated prior estimates of disease probability with clinical evidence from literature-reported phenotyped individuals, arrhythmia genetics clinical cohorts, and population controls, refined by variant-specific features including structural, *in silico* variant classifiers (AlphaMissense), and variant MAVE scores reported here. These prior estimates of LQTS penetrance conditioned on variant-specific features were then integrated with previous observations and clinical diagnoses of individuals heterozygous for *KCNH2* variants, affected (diagnosed) or unaffected (or reasonably estimated not to have the disease) through a beta-binomial Bayesian model; affected and unaffected heterozygote counts come from the literature (removing all potential overlapping individuals in the curated clinical and UK Biobank cohorts) and gnomAD v4.0.0. The outcome of this comprehensive analysis is a set of posterior penetrance estimates, which are available for community use via variantbrowser.org^6,13,14^.

**Clinical Analyses**

*Clinical Phenotyping from Variant Heterozygotes.* For our study, we gathered clinical phenotypes and genotypes for *KCNH2* probands and their relatives from four international arrhythmia genetics centers located in the USA, Italy, Japan, and New Zealand. Each center, recognized for its expertise in this field, received approval from local institutional review boards or ethics committees. They conducted a retrospective analysis of patient records, ensuring anonymity of patient identifiers in line with previously established protocols^6,13^. UK Biobank was accessed through the Online Research Access Platform, under Application ID 94960. Using the UKB cohort browser, demographic, medical, and genetic data from all 502,364 individuals were collected. Demographic data included their de-identified ID, year of birth, sex (self-reported), age at recruitment, and ethnicity (self reported). Medical data included QT intervals, raw and corrected, age at ECG, history of defibrillation, syncope, hypertrophic cardiomyopathy, cardiac arrest, atrial fibrillation, atrial flutter, ventricular fibrillation or flutter, medication list, BMI, and ICD status. Genetic information included presence and burden of SNP variants in *KCNH2*. For patients with multiple quantitative measurements of the same variable at different time points, the earliest available measurement was used. The final dataset was downloaded using the Table Browser function of the UKB RAP.

*Feature selection and time-to-event model*. To address feature multicollinearity (Supplemental Figure 5) and ensure parsimonious, robust time-to-event models of cardiac event outcomes, we used Variance Inflation Factor (VIF) and Least Absolute Shrinkage and Selection Operator (LASSO) within a Cox Proportional Hazards (CoxPH) model to identify and validate significant, variant-specific covariates. Both VIF and LASSO CoxPH were calculated using the variant-specific covariates of APC peak tail current, MAVE trafficking score, AlphaMissense, LQTS penetrance estimate (incorporating all prospectively available variant-specific features and affected/unaffected heterozygotes distinct from the clinical cohort presented here), ClinVar annotation (with VUS and Conflicting set to ‘0’, or ‘Benign’), and long QT syndrome structure-derived penetrance estimate^15^. The covariates with VIF values > 5 were removed (only the structurally derived LQTS penetrance estimate met this criterion). We then conducted a sex-stratified Cox PH survival analysis, incorporating LASSO, to the time first cardiac event: syncope, VT, VF, aborted cardiac arrest, or sudden unexplained death (Supplemental Figure 7); tested features and resulting models satisfied the proportional hazards assumption at the 0.01 significance level; for this step, all covariates were scaled to a mean of 0 and standard deviation of 1. We identified corrected QT interval, APC peak tail current, and LQTS penetrance as the most informative covariates; MAVE score was insignificant at all regularization penalty thresholds (see also Figure 6). With this reduced feature list, we built parametric Royston-Parmar survival models again with an age cutoff at 40 years using the `flexsurvspline` function from the R package `flexsurv`; hazard ratio estimates and 95% confidence intervals of resulting multivariate models were generated via bootstrap with resampling 500 times. To visualize time-dependent ROC curves from the Royston-Parmar survival models, we used a nearest neighbor estimation for censored data the ‘survivalROC’ package in R and a target age of 20 years old^16^.

**Supplemental Discussion**

The MAVE and APC assays are calibrated to detect different instances of K_v_11.1 dysfunction. In human cardiomyocytes, K_V_11.1 is a tetrameric channel subject to dominant negative effects between a WT copy and certain loss-of-function variant copies. The MAVE trafficking assay is a ‘hemizygous’ assay, in which a single copy of Kv11.1 is present in each cell. In contrast, the APC assay uses an IRES-based plasmid to incorporate a single unique combination of WT and variant *KCNH2* per cell. Therefore, completely haploinsufficient variants would be recorded as 0% WT trafficking in the MAVE, while recording 50% peak tail current in the APC assay. Furthermore, the APC will detect not only trafficking deficient variants (type 2 defect), but also variants that affect channel gating (Class 3 defect) and potassium ion permeability (Class 4 defect)^17^.

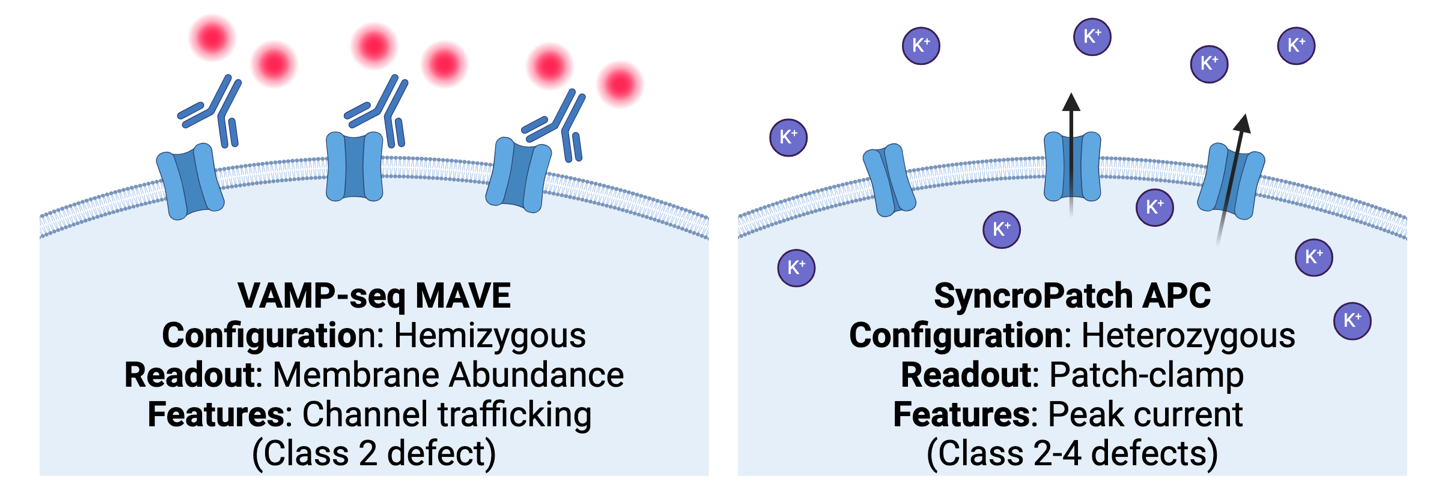

For both assays, the functional scores are best calibrated to detect loss-of-function versus gain-of-function variants. Gain-of-function variants in *KCNH2* and other cardiac ion channels are associated with an extremely rare condition, Short QT Syndrome (OMIM: 609620). We do not attempt to detect these variants here. For normalization of functional data as input to clinical analyses, we capped all functional scores at 100% of WT.

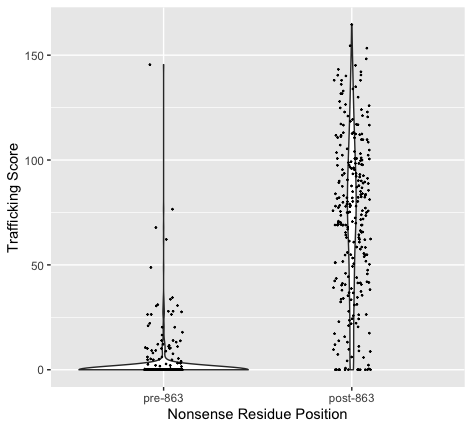

**Supplemental Figure 1. Trafficking Scores of Nonsense Residues Before and After Residue 863.** The mean trafficking score of nonsense variants up to residue 863 was 1.35 ± 7.86, and after 863 it was 71.7 ± 39.1.

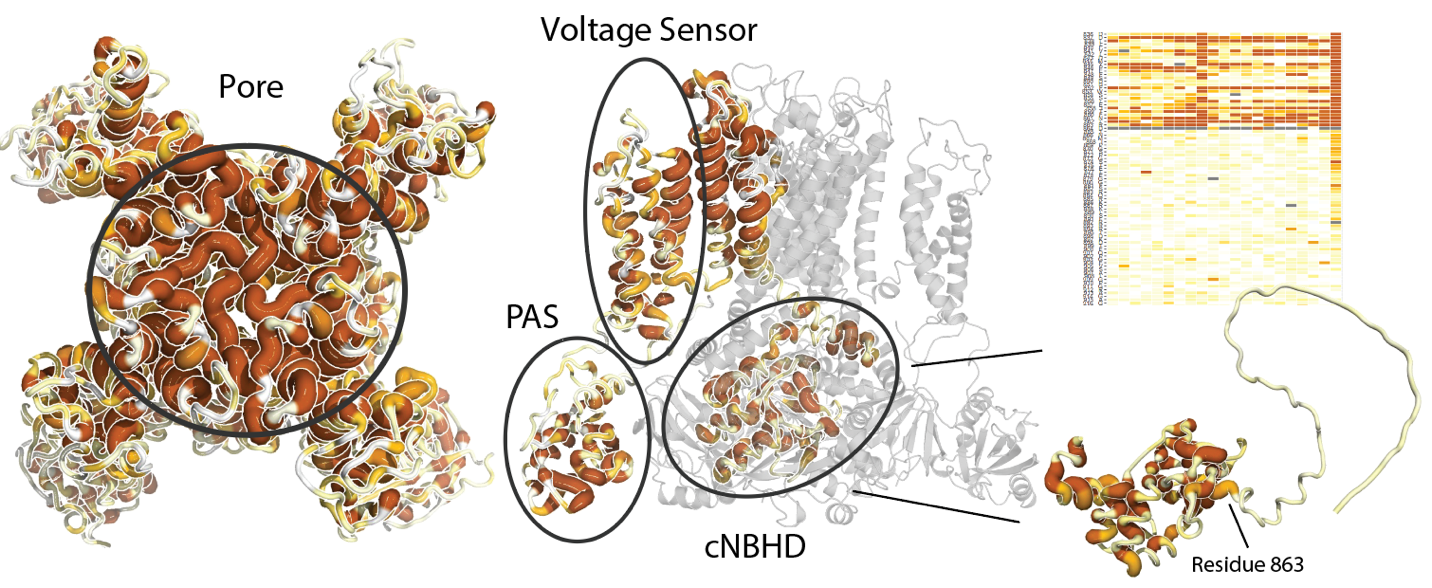

**Supplemental Figure 2. Structural Heterogeneity of “hot spot” Domains.** Heterogeneity of MAVE scores in *KCNH2* “hot spot” domains imposed on channel structure. Cartoon thickness and color correspond with per-residue average trafficking scores. PAS (Per-Arnt-Sim) domain, cNBHD (cyclic-Nucleotide Binding Homology Domain).

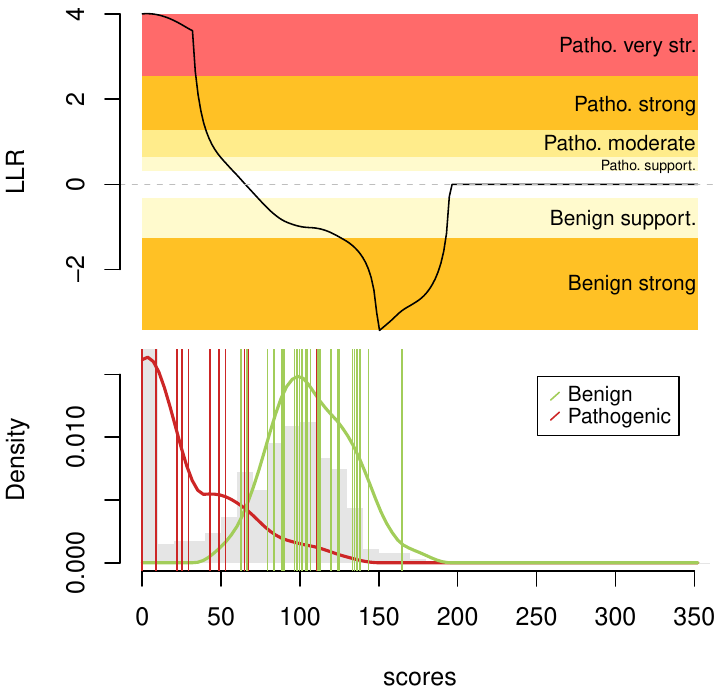

**Supplemental Figure 3.** Log-likelihood ratios derivation for MAVE.

Individual red and green vertical lines indicate the control pathogenic and benign variants, respectively, used to calibrate the assay.

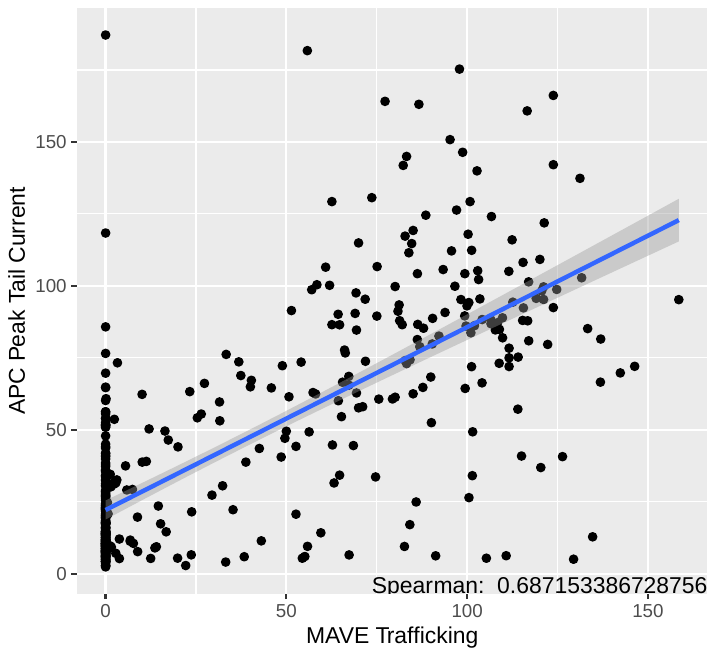

**Supplemental Figure 4. Correlation of MAVE and APC Functional Scores.** Spearman rank-order correlation (0.69) of MAVE and APC raw functional scores for 443 variants studies by both methods.

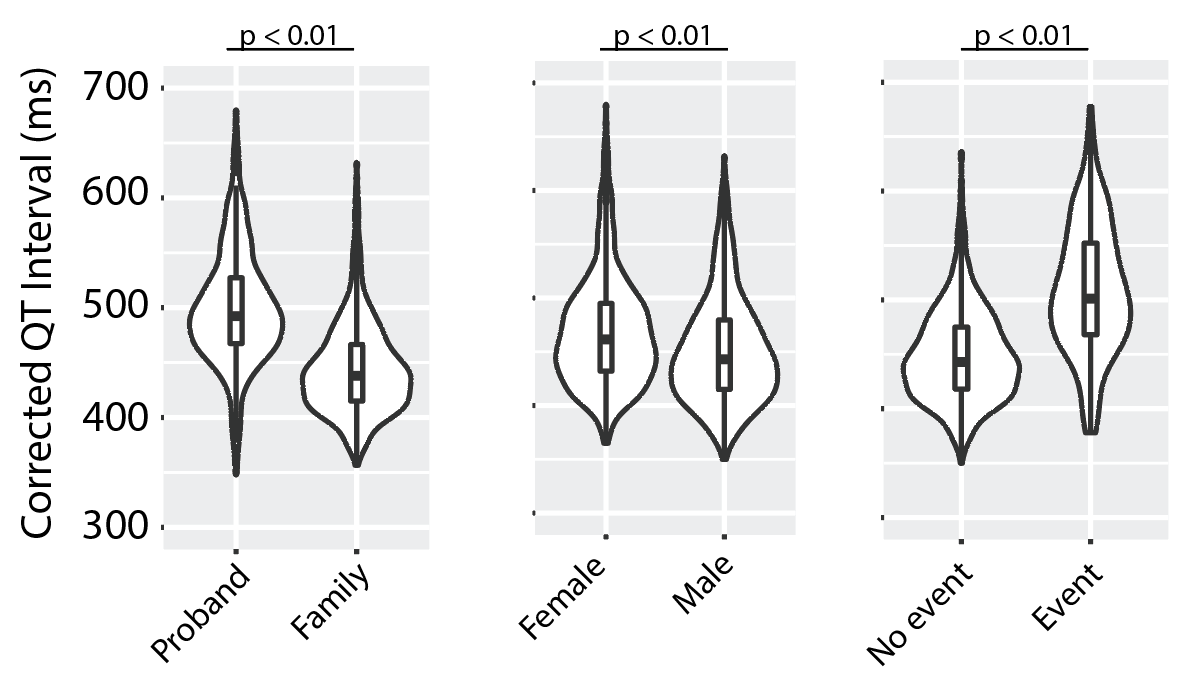

**Supplemental Figure 5.** Plots of cohort corrected QTc intervals.

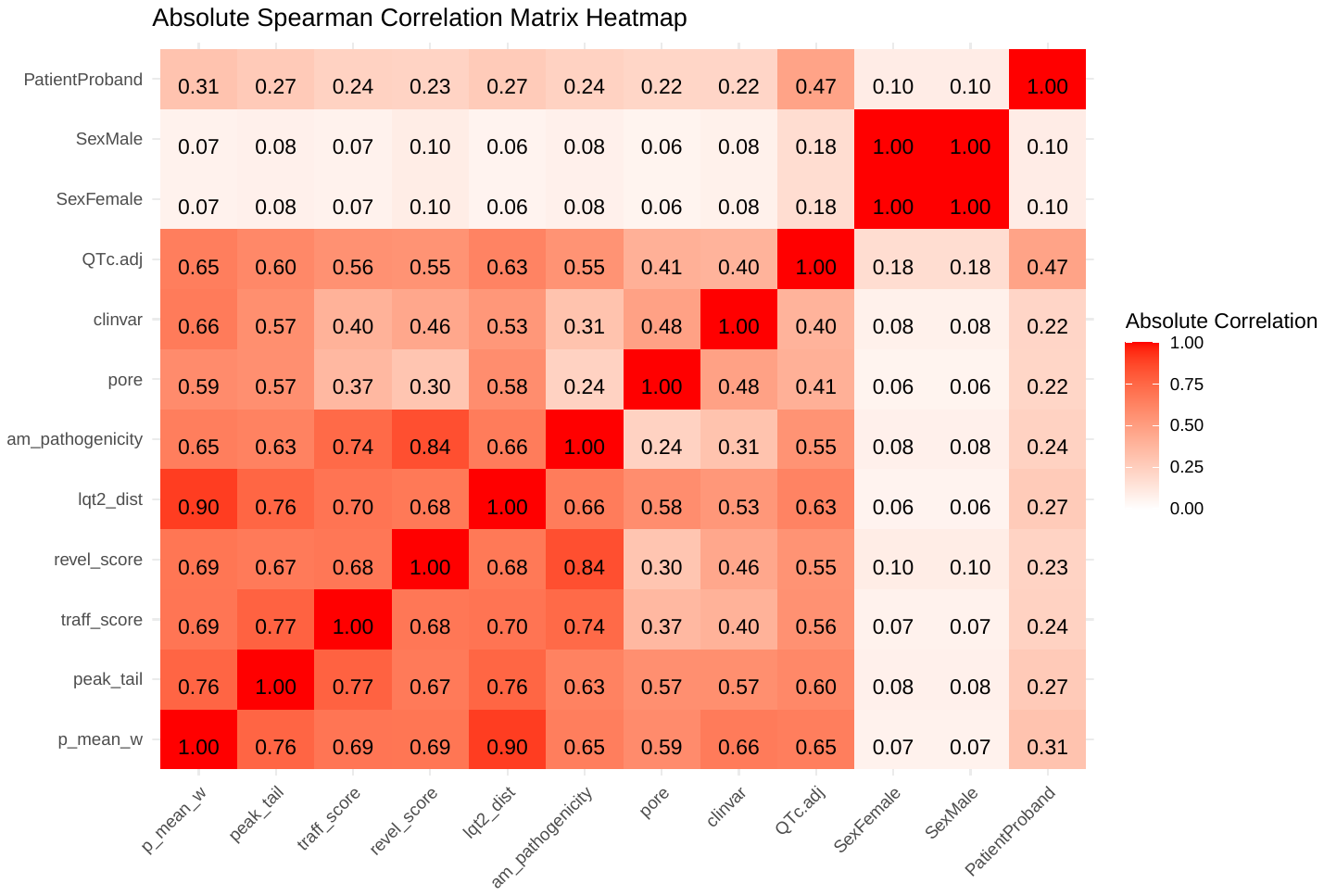

**Supplemental Figure 6.** Co-linearity analysis of all model covariates considered by Spearman correlation analysis.

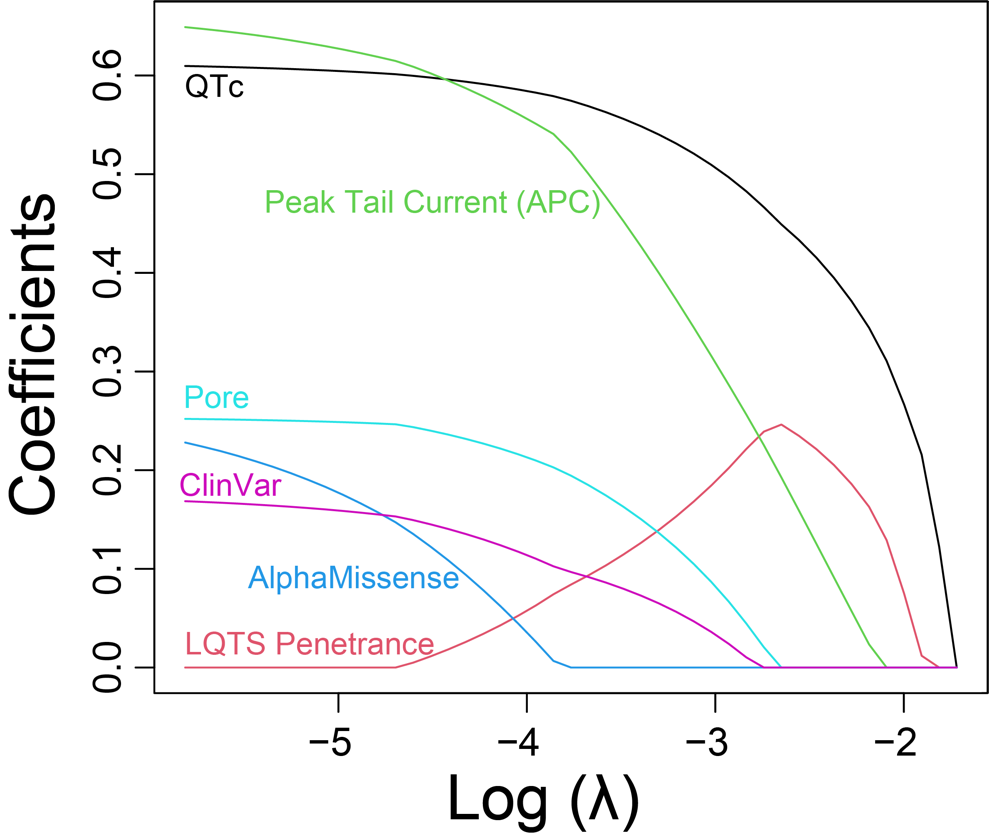

**Supplemental Figure 7. Regularization path analysis for a** **Least Absolute Shrinkage and Selection Operator (LASSO) within a Cox Proportional Hazards time-to-event model.** The coefficients of the predictors in the time-to-event, sex stratified, Cox proportional hazards model mentioned above as a function of the log-transformed λ value, the regularization parameter. The paths demonstrate each covariate's coefficient in the model as a greater penalty is applied, increasing λ. The x-axis shows the log-transformed values of λ, and the y-axis shows the coefficients of the covariates in the Cox model.
